# Knowledge on tuberculosis, services of TB control programme and associated socio-demographic inequity among rural participants of Jaipur, Rajasthan, India: A cross sectional study

**DOI:** 10.1101/2022.12.19.22283674

**Authors:** P.K. Anand, V. Dhikav, M. Rathore, G.S. Toteja, Mradula Singh, Bhanwar Manohar Singh, C.R. Meena, Anil Jangid, M.R. Devanda, M.L. Mathur

**Affiliations:** ICMR – National Institute for Implementation Research on Non Communicable Diseases, New Pali Road, Jodhpur, Rajasthan, India – 342005; Model Rural Health Research Unit, Department of Health Research, Government of India, Bhanpur Kalan, Jaipur, Rajasthan, India – 302028; Department of Preventive & Social Medicine, Sawai Man Singh Medical College & associated hospitals, Jaipur, Rajasthan, India – 302004

**Keywords:** Knowledge, Knowledge, Tuberculosis, Inequity, India

## Abstract

**Introduction:** Tuberculosis (TB) is one of the top 10 causes of deaths worldwide. India is still a highest TB burden country. There is scarcity of data on TB knowledge from Rajasthan state of India.

**Materials & Methods:** Cross-sectional community based study was carried out at Model Rural Health Research Unit, Jaipur, an unit of Department of Health Research, Ministry of Health & Family Welfare, Government of India.

**Results:** Study reports the result from 1993 adult participants from 10 villages of 2 sub-districts of district Jaipur. About 88.9% of studied participants knew that TB is an infectious disease and it spreads from TB patient to healthy person in close contact. Only 22.3% of participants knew ‘DOTS is the name of treatment for TB’. While, only 58.9% knew ‘*sputum is used for diagnosis of TB*’ at health centres. *Scheduled castes, scheduled tribes* and *backward classes* social groups knew less than the mainstream ‘*General*’ social group. The observed difference was statistically significant (p<0.05). Logistic regression analysis estimated the relative contribution in knowledge status.

**Discussion:** The knowledge of study participants on transmission of tuberculosis was similar to the knowledge of population in country wide study. They poorly knew that sputum is used for diagnosing the tuberculosis disease; socio-demographic inequity exists in this knowledge too. People from older age groups, underprivileged social groups and minority need extra educational activities.

## Introduction

Tuberculosis (TB) is one of the top 10 causes of deaths worldwide. Eight countries of the world, which includes India, account for two thirds of global total of TB cases [1]. The Global Plan and Stop TB Partnership 2006-2015 visualized a TB free world. To ensure the TB free world, access to effective diagnosis, treatment and cure by every TB patient is one among five missions of Global Plan. India is still a highest TB burden country globally in regards of absolute numbers of incident cases each year. Mortality due to this disease is the third leading cause of years of life lost (YLLs), in India [2].

Lack of knowledge on tuberculosis has been found to be associated not only with occurrence of tuberculosis [3] but also with delayed diagnosis [4-7].

Cross sectional studies on tuberculosis knowledge have been published for different parts of India [8-20]. Nationwide studies for India were conducted on secondary data and are more than a decade old [13, 16].

Rajasthan state of India is located in the north-western part of the country. It shares state boundary on the west and northwest with Pakistan, on the north and northeast with states of Punjab, Haryana, and Uttar Pradesh, on the east and southeast with states of Uttar Pradesh and Madhya Pradesh, and on the southwest with Gujarat. The capital city is Jaipur [21].

Recently conducted study by ICMR on prevalence of tuberculosis in India reports prevalence for the country and its states separately. The prevalence of microbiologically confirmed tuberculosis among population aged > 15 years were 316 and 484 per 100 000 population covered for India and Rajasthan state. Rajasthan state of India is having more prevalent cases of TB then the country [22].

There is scarcity of available published articles on TB knowledge from Rajasthan state in general and Jaipur district in particular. The published studies available from Rajasthan were conducted on small sample sizes in other places of state [23-24].

Therefore, this study was carried out to estimate the prevalence of knowledge about TB and services of TB control programme and to determine its correlates among rural population of selected villages of Jaipur, hitherto unreported area.

## Material & Methods

### Study Design

Cross-sectional community based study

### Study setting

This study was carried out among rural participants aged ≥ 15 years at Model Rural Health Research Unit, Jaipur, a unit of Department of Health Research, Ministry of Health & Family Welfare, Government of India during August 2014 till July 2017.

### Study participants

Individuals of age ≥ 15 years residing in selected study villages from both the genders were our study participants.

### Sample size

Sample size of rural participants aged ≥ 15 years was calculated by assuming expected prevalence (p) of knowledge of tuberculosis among 50.0%; 95% confidence limits; and 2% absolute precision error. We calculated sample size of 2395 in Epi-info version 7.2.4.0 [25].

### Sampling Design

Two stage simple random sampling design was adopted. In first stage, 2 sub-districts/tehsils namely Jamwa Ramgarh and Sanganer were selected from the district Jaipur. In second stage, 5 villages were randomly selected from each of the selected 2 sub-districts/tehsils of district Jaipur. From each of the 10 study villages, 100 households were selected. Household in the central location in village was first of all selected. From this selected central Household all consecutive households in East direction were chosen. In case of non availability of consecutive households in east direction, then further selection were made of consecutive households in west direction of selected central household. Similarly we repeated this strategy for north and south direction till desired number of households were chosen from each village. From each of the selected household, each of the available individuals aged ≥ 15 years available at the time of interview were registered in the study after seeking consent.

### Data collection team

A data collecting team was comprised involving scientists, and technical staffs posted at Model Rural Health Research Unit. All team members were discussed about the study, its objectives, & methodologies. They were trained through lecture and demonstration in seeking consent from participants, administration of study questionnaire, and collection of data. Questions asked by the team members during training were answered by principal investigator and resolved up to the satisfaction level of members of ‘Data collection team’.

### Study questionnaire

Study questionnaire was developed in English language by reviewing previous published reports & research papers and having discussion among study investigators. Study questionnaire had broadly three parts, (a) participant’s information and his/her socio-demographic details; (b) tuberculosis and anti-tubarcular treatment in family and (c) questions with regard to knowledge on tuberculosis and services of TB control programme. Study questionnaire was then translated in to vernacular language ‘Hindi’. ‘Hindi’ questionnaire was then back translated in to English questionnaire. These questionnaires were checked for content consistency. Hindi questionnaire meeting content consistency with English questionnaire was applied to members of data collection team to ensure its comprehensibility. After this comprehensibility testing among team members, questionnaire was applied to sample participants in study village. Corrections were made in the questionnaire as per the understanding of study participants. After this testing of questionnaire, finally developed questionnaire was applied to entire study sample. Final questionnaire comprised of 15 variables in total in its 3 parts including (a) participant’s information and his/her socio-demographic details, (b) History of tuberculosis and antitubercular treatment in family, and (c) questions with regard to knowledge on tuberculosis and services of TB control programme. Sample participants under testing of questionnaire was not part of study sample. Study questionnaire was administered to the selected study participants through face-to-face interview by the trained interviewer who was a member of the ‘data collection team’. Obtained responses of the study participants were recorded in study questionnaire by interviewer.

#### Study variables

Study variables chosen after reviewing previous published research papers and reports were classified in following categories:-

##### 1) Outcome variables

Study used total 6 categorical variables as outcome variables. These variables were on knowledge of tuberculosis and services of TB control programme. Questions framed to elicit information on these outcome variables were:-

a. Do you know TB spreads from its patient to healthy person in contact?
b. Do you know the way tuberculosis spreads?
c. Is TB treatable?
d. DOTS is the name of treatment for which disease?
e. Do you know diagnosis and treatment of TB are provided completely free of cost at government health centres?
f. How TB is diagnosed at government health centre?

##### 2) Predictor variables

Study used total 4 predictor variables as socio-demographic predictor variables to assess the association if any of outcome variables. These predictor variables were (1) age, (2) gender, (3) religion and (4) Social group.

###### Social group

Data pertaining to the name of the social group, known locally as *caste*, of individual participant was collected as per the Government of Rajasthan guidelines. On the basis of prevalence of illiteracy, participation in government owned services, and perceived social status there are 5 social groups. These characteristics are the basis for deciding on the caste based government sponsored reservation system for community upliftment in Rajasthan. These social groups are *scheduled caste* (*SC*), *scheduled tribe* (*ST*), *other backward castes* (*OBC*), m*ost backward castes* (*MBC*), and *general* social group. *General* social group of India is the main stream social group and it was considered as the yardstick for comparing other social groups [26-27]. Data on social group of individual study participant was recorded as reported by self.

##### 3) Confounding variables

Following variables were chosen as confounding variables, because these could bias the result. The considered confounding variables were (1) History of tuberculosis disease or anti-tubercular therapy among any family member of study participant, (2) History of tuberculosis disease or anti-tubercular therapy among study participant, (3) Any family member including study participant receiving anti-tubercular treatment in present days.

### Statistical analysis

Data was collected by administering pretested questionnaire. Data collected on paper based questionnaire was then entered in Microsoft Excel computer programme through double data entry. Both files of entered study data were then compared in Epi-info for assessing accuracy. Accurate data was analyzed in statistical software. Affirmative responses with ‘confounding variables’ were filtered out from statistical analysis. Studied outcome variables and predictor variables were summarized as (%) proportions in table. Outcome variables with knowledge among less than 60% participants were chosen arbitrarily for computing bivariate and multivariable logistic regression analysis. Considered outcome variables were dichotomized for performing bivariate analysis & multivariable logistic regression analysis with considered categories of predictor variables. We used χ^2^ as the test of difference in proportions in bivariate analysis to detect the associated predictor. Bivariate analysis was performed in 2×2 table function of Epi-info 7. Since there were more than 1 type of predictor variables in study to analyze their association with considered dichotomized outcome variable, multivariable logistic regression analysis was performed in Epi-info 7 to discern the independently associated predictor variable out of all the predictor variables fitted in model. This analysis was performed separately with each of the considered dichotomized outcome variables. Adjusted Odds Ratio was estimated under multivariable logistic regression analysis to identify the independently associated predictor variable and p value less than 0.05 was considered statistically significant. Analysis was carried out in Epi-info 7 version 7.2.4.0 [25].

### Ethical conduct

study was evaluated by Institute Ethics Committee (IEC) of ICMR – National Institute for Implementation Research on Non Communicable Diseases, Jodhpur. This study got approval by IEC. Informed & written consent was sought from the study participants by a member of the ‘Data collection team’ other than the interviewer. Study was carried out following standard ethics procedures [28].

### Study reporting

Study was carry out as per the STROBE guidelines [29].

## Results

We recruited 2293 participants in the study. Participants with past history or current suffering with tuberculosis and/or in their family members were removed from data, as they could bias the knowledge status of general population and skewed the sample statistic. Participant’s data with ‘missing’ information on one or few variables was removed (n=300). Finally 1993 participant’s data with information on all variables were subjected for statistical analysis.

Table 1 describes the knowledge on tuberculosis and services of National TB elimination programme among the studied participants. About 88.9% of studied participants knew that TB is an infectious disease and it spreads from TB patient to healthy person in close contact. Knowledge on mode of transmission of tuberculosis was adequate. About 88.9% of studied participants knew that the disease spreads through coughing or sneezing by TB patient to healthy person in close contact. About 80.3% of participants knew that TB is treatable. About 72.5% of participants knew that the diagnosis and treatment of TB are provided free of cost at government run health centres. Only 22.3% of participants knew that ‘DOTS is the name of treatment for TB’. Only 58.9% knew that the diagnosis of TB is made through examining the sputum at health centres.

**Table 1:**
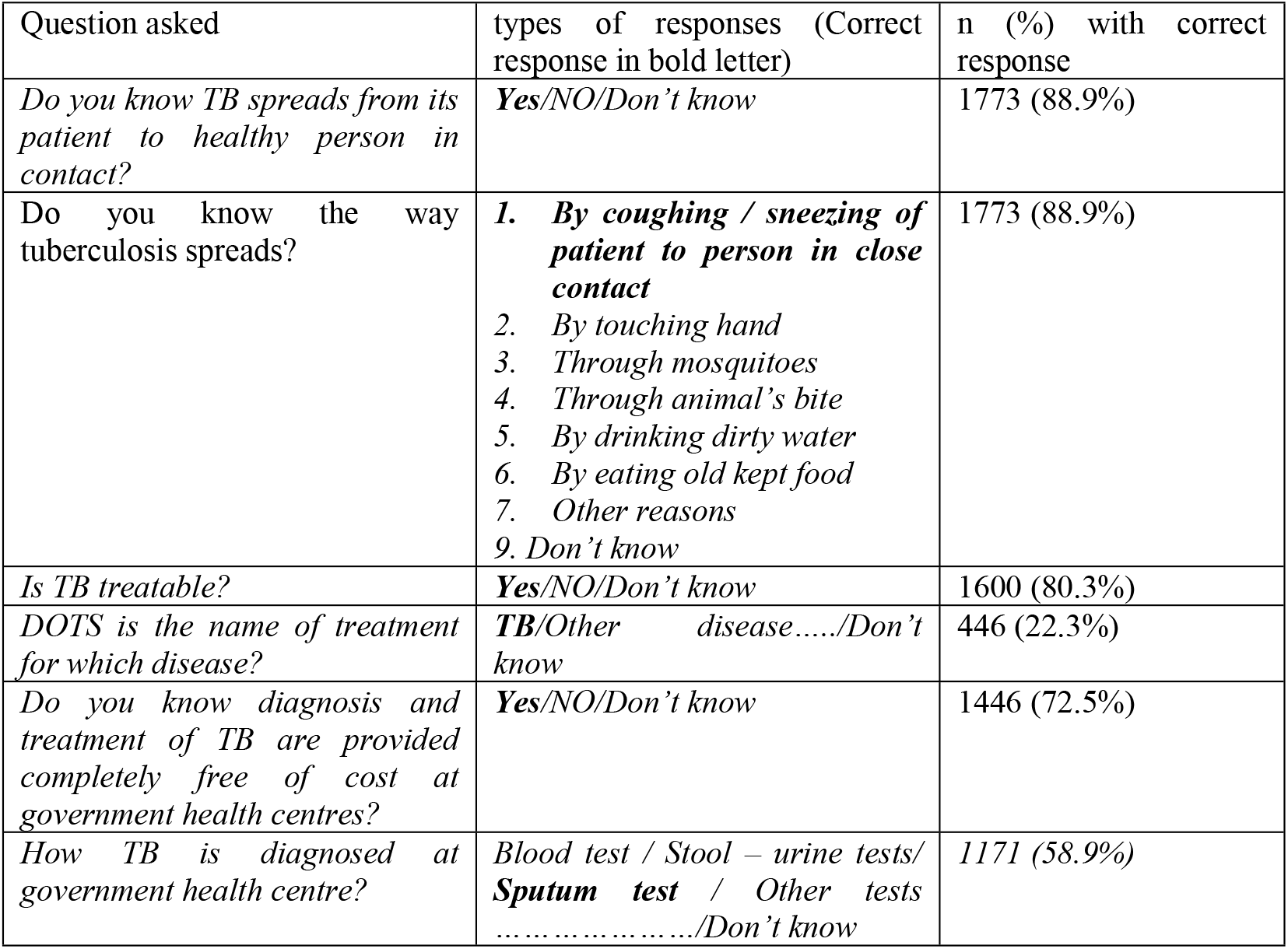
Knowledge on tuberculosis and services of TB control programme among study participants (n=1993)

| Question asked | types of responses (Correct response in bold letter) | n (%) with correct response |
| --- | --- | --- |
| *Do you know TB spreads from its patient to healthy person in contact?* | **Yes**/NO/Don't know | 1773 (88.9%) |
| Do you know the way tuberculosis spreads? | **1. By coughing / sneezing of patient to person in close contact**<br>2. By touching hand<br>3. Through mosquitoes<br>4. Through animal's bite<br>5. By drinking dirty water<br>6. By eating old kept food<br>7. Other reasons<br>9. Don't know | 1773 (88.9%) |
| *Is TB treatable?* | **Yes**/NO/Don't know | 1600 (80.3%) |
| *DOTS is the name of treatment for which disease?* | **TB**/Other disease...../Don't know | 446 (22.3%) |
| *Do you know diagnosis and treatment of TB are provided completely free of cost at government health centres?* | **Yes**/NO/Don't know | 1446 (72.5%) |
| *How TB is diagnosed at government health centre?* | Blood test / Stool – urine tests/<br>**Sputum test** / Other tests<br>...../Don't know | 1171 (58.9%) |

Table 2 details on the knowledge on ‘DOTS is the name of treatment for TB’ among studied socio-demographic factors. It analyzed the differences in proportions of participants between who know and who don’t know ‘DOTS is the name of treatment for TB’ with respect to their socio-demographic factors. Overall the knowledge on this parameter was low across all the studied factors. None of the type of any socio-demographic factor reached knowledge in 50.0% participants. The proportions of participants from different social groups were compared with ‘*General*’ caste group as benchmark. As Table 2 depicts 34.5% ‘*General*’ social group participants knew ‘DOTS is the name of treatment for TB’. The proportions of participants belonging to ‘*SC*’, ‘*MBC*’, & ‘*OBC*’ social groups who knew ‘DOTS is the name of treatment for TB’ were 10.1%, 18.3% & 25.3% respectively. All these three social group participants were statistically significantly knowing less ‘DOTS is the name of treatment for TB’ than ‘*General*’ social group as benchmark (p<0.05). The proportion of *ST* participants who knew ‘DOTS is the name of treatment for TB’ was 28.1%. The proportion of *ST* participants was less compared to proportion of ‘*General*’ social group, though the difference was statistically insignificant. Similarly, proportions of participants who know and who don’t know ‘DOTS is the name of treatment for TB’ with respect to their age groups were compared. The age group 15 – 24 years age group was used as benchmark. About 33.8% of participants of age group 15 – 24 years age group knew ‘DOTS is the name of treatment for TB’. The proportions of participants belonging to age groups 25 – 39 yrs, 40 – 59 yrs and 60 yrs who knew ‘DOTS is the name of treatment for TB’ were 21.2%, 15.2% and 8.8% respectively. These age groups participants had statistically significantly low knowledge than the 15 – 24 yrs age group (p<0.05). About 25.3% female knew compared to 19.8% of male about this parameter. The proportion of females who knew was significantly more than the proportion of males who knew (p<0.05). Knowledge on ‘DOTS is the name of treatment for TB’ did not differ significantly with respect to religion.

**Table 2:** Knowledge on ‘DOTS is the name of treatment for TB’ among studied socio-demographic factors of participants (n=1993)

| Socio-demographic factors | Sub-group | Knowledge on 'DOTS is the name of treatment for TB' |  |  |  |
| --- | --- | --- | --- | --- | --- |
|  |  | Know | Don't know | X<sup>2</sup> value | p value |
| Social Groups* | ST(n=444) | 125 (28.1%) | 319 (71.9%) | 2.65 | 0.10 |
|  | SC(n=445) | 45(10.1%) | 400 (89.9%) | 56.7 | **0.0001** |
|  | MBC(n=316) | 58 (18.3%) | 258 (81.7%) | 17.3 | **0.0001** |
|  | OBC(n=585) | 148 (25.3%) | 437 (74.7%) | 6.3 | **0.01** |
|  | General(n=203) | 70 (34.5%) | 133(65.5%) | reference | reference |
| Age groups | ≥60 Years(n=91) | 8 (8.8%) | 83 (91.2%) | 23.0 | **0.0000** |
|  | 40 – 59 years(n=453) | 69 (15.2%) | 384 (84.8%) | 43.8 | **0.0000** |
|  | 25 – 39 years(n=956) | 202 (21.2%) | 754 (78.8%) | 27.8 | **0.0000** |
|  | 15 – 24 years(n=493) | 167 (33.8%) | 326 (66.2%) | reference | reference |
| Gender | Female (n=932) | 236 (25.3%) | 696 (74.7%) | 8.73 | **0.003** |
|  | Male (n=1061) | 210 (19.8%) | 851 (80.2%) | reference | reference |
| Religion | Hindu (n=1974) | 442 (22.4%) | 1532 (77.6%) |  | 1.0000** |
|  | Muslim (n=19) | 4 (21.0%) | 15 (79.0%) |  | reference |

\*SC=Scheduled caste; ST=Scheduled tribe; MBC=Most Backward caste; OBC=other backward caste
\*\* Fisher exact test 2 tailed p value

Table 3 details knowledge on ‘*sputum is used for diagnosis of TB’* among studied socio-demographic factors. The differences in proportions of participants who know and who don’t know ‘*sputum is used for diagnosis of TB’* with respect to studied socio-demographic factors on are also given. As Table 3 depicts 72.4% ‘*General*’ social group participants knew ‘*sputum is used for diagnosis of TB*’. The proportions of participants belonging to *ST, SC*, & ‘OBC’ social groups who knew ‘*sputum is used for diagnosis of TB*’ were 61.3%, 49.6% & 54.0% respectively. All these three social group participants were statistically significantly less knowing ‘*sputum is used for diagnosis of TB*’ than the ‘*General*’ social group as benchmark (p<0.05). The proportion of *MBC* participants knowing ‘*sputum is used for diagnosis of TB*’ was 69.0%. These *MBC* participants knew less compared to ‘*General*’ social group, though the difference was statistically insignificant.

**Table 3:** Knowledge on ‘*sputum is used for diagnosis of TB’* among studied socio-demographic factors of participants (n=1993)

| Socio-demographic factors | Sub-group | Knowledge on '*sputum is used for diagnosis of TB*' |  |  |  |
| --- | --- | --- | --- | --- | --- |
|  |  | Know | Don't know | X<sup>2</sup> value | p value |
| Social groups* | *ST* (n=444) | 272 (61.3%) | 172 (38.7%) | 7.59 | **0.005** |
|  | *SC* (n=445) | 221 (49.6%) | 224 (50.4%) | 29.4 | **0.0001** |
|  | *MBC* (n=316) | 218 (69.0%) | 98 (31.0%) | 0.69 | 0.4 |
|  | *OBC* (n=585) | 316 (54.0%) | 269 (46.0%) | 21.0 | **0.0001** |
|  | *General* (n=203) | 147 (72.4%) | 56 (27.6%) | reference | reference |
| Age groups | ≥60 Years (n=91) | 48 (52.7%) | 43 (47.3%) | 3.46 | 0.06 |
|  | 40 – 59 years (n=453) | 237 (52.3%) | 216 (47.7%) | 11.2 | **0.0008** |
|  | 25 – 39 years (n=956) | 578 (60.4%) | 378 (39.6%) | 0.94 | 0.33 |
|  | 15 – 24 years (n=493) | 311 (63.0%) | 182 (37.0%) | reference | reference |
| Gender | Female (n=932) | 531 (56.9%) | 401 (43.1%) | 2.69 | 0.10 |
|  | Male (n=1061) | 643 (60.6%) | 418 (39.4%) | reference | reference |
| Religion | Hindu (n=1974) | 1169 (59.2%) | 805 (40.8%) | 8.4 | **0.003** |
|  | Muslim (n=19) | 5 (26.3%) | 14 (73.7%) | reference | reference |

\*SC=Scheduled caste; ST=Scheduled tribe; MBC=Most Backward caste; OBC=Other backward caste

Similarly, proportions of participants who know and who don’t know ‘*sputum is used for diagnosis of TB*’ with respect to their age groups were compared. The age group 15 – 24 years age group was used as benchmark. About 63.0% of participants of age group 15 – 24 years age group knew ‘*sputum is used for diagnosis of TB*’. The proportion of participants belonging to age groups 25 – 39 yrs, 40 – 59 yrs and 60 yrs and above who knew ‘*sputum is used for diagnosis of TB*’ were 60.4%, 52.3% and 52.7% respectively. Though the participants of these age groups knew less than the 15 – 24 yrs age group, the difference was statistically significant in case of 40 – 59 yrs age group only (p<0.05). About 56.9% females knew compared to 60.6% males about this parameter. Though the proportion of females who knew was less, however the observed difference was statistically not significant. About 59.2% of Hindus knew compared to 26.3% Muslims about this parameter. The observed difference in proportions was statistically significant (p<0.05).

Table 4 details on the results of logistic regression analysis for determining the relative contribution of studied socio-demographic factors in knowledge on ‘DOTS is the name of treatment for TB’ among studied participants. Odds of occurrence of knowledge on ‘DOTS is the name of treatment for TB’ among age groups 25 – 39 yrs, 40 – 59 yrs, and 60 yrs and above were 0.49, 0.32 and 0.18 against the age group 15 – 24 yrs. The observed lesser odds of occurrence was statistically significant independently in these age groups (p<0.05). Males had 0.71 Odds of occurrence of knowledge on ‘DOTS is the name of treatment for TB’ as compared to females, it was statistically significant (p<0.05). Considered social groups viz. *MBC, OBC, SC* & *ST* had 0.31, 0.56, 0.17 & 0.65 Odds ratio against *General* social group. The values of odds ratio were statistically significantly lower among these social groups compared to *General* social group (p<0.05).

**Table 4:** Multivariable logistic regression analysis of socio-demographic factors with knowledge on ‘*DOTS is the name of treatment for TB’* (n=1993)

| socio-demographic factors | Adjusted Odds ratio | 95% Confidence interval | p value |
| --- | --- | --- | --- |
| Age group (25 – 39 yrs/15 – 24 yrs) | 0.49 | 0.38 – 0.63 | **0.0000** |
| Age group (40 – 59 yrs/15 – 24 yrs) | 0.32 | 0.23 – 0.45 | **0.0000** |
| Age group (60 yrs & above/15 – 24 yrs) | 0.18 | 0.08 – 0.39 | **0.0000** |
| Religion (Muslim/Hindu) | 0.83 | 0.26 – 2.6 | 0.76 |
| Sex (Male/Female) | 0.71 | 0.56 – 0.89 | **0.003** |
| Social Group (MBC/General) | 0.31 | 0.20 – 0.47 | **0.0000** |
| Social Group (OBC/General) | 0.56 | 0.39 – 0.80 | **0.0015** |
| Social Group (SC/General) | 0.17 | 0.11 – 0.26 | **0.0000** |
| Social Group (ST/General) | 0.65 | 0.45 – 0.93 | **0.0218** |

Table 5 details on the results of logistic regression analysis for determining the relative contribution of studied socio-demographic factors in knowledge on ‘*sputum is used for diagnosis of TB*’ among studied participants. Odds of occurrence of knowledge among 40 – 59 yrs and 60 yrs and more were 0.6 in both age groups compared to 15 – 24 yrs age group. The observed value of odds ratio was statistically significant at p<0.05. Muslim had 0.25 odds ratio compared with Hindu and the difference was statistically significant (p<0.05). Males had odds ratio of 1.29, which was statistically significantly higher than females (p<0.05). Social groups viz. *OBC, SC* & *ST* had Odds ratios of 0.46, 0.37 & 0.6 respectively, and these were statistically significantly lower than the compared *General* social group (p<0.05).

**Table 5:** Multivariable logistic regression analysis of socio-demographic factors with knowledge on ‘*sputum is used for diagnosis of TB’* among studied participants (n=1993)

| socio-demographic factors | Adjusted Odds ratio | 95% Confidence interval | p value |
| --- | --- | --- | --- |
| Age group (25 – 39 yrs/15 – 24 yrs) | 0.89 | 0.71 – 1.13 | 0.36 |
| Age group (40 – 59 yrs/15 – 24 yrs) | 0.60 | 0.46 – 0.78 | **0.0002** |
| Age group (60 yrs & above/15 – 24 yrs) | 0.60 | 0.38 – 0.96 | 0.0333 |
| Religion (Muslim/Hindu) | 0.25 | 0.08 – 0.71 | 0.0097 |
| Sex (Male/Female) | 1.29 | 1.07 – 1.56 | **0.0073** |
| Social Group (MBC/General) | 0.88 | 0.59 – 1.31 | 0.53 |
| Social Group (OBC/General) | 0.46 | 0.33 – 0.66 | **0.0000** |
| Social Group (SC/General) | 0.37 | 0.25 – 0.53 | **0.0000** |
| Social Group (ST/General) | 0.60 | 0.41 – 0.86 | **0.0062** |

## Discussions

Tuberculosis is an infectious disease caused by *Mycobacterium tuberculosis* bacterium. The disease transmits from infective patient to healthy person in close contact through micro droplets formed during coughing and sneezing. In our study, 88.9% of participants knew that TB is an infectious disease which transmits from its patient to healthy person in close contact.

Previous population based studies reported more than 80.0% knew TB is an infectious disease [9] [16]. There are some previous studies which reported less than 80.0% knew TB is an infectious disease also. Study by Kulkarni P *et al* carried out among tribal population of Jharkhand reported 61.0% participants knew TB as a transmissible disease [10]. Another cross sectional study conducted among 360 college students of Karnataka showed 56.7% knew TB spreads from person to person [17]. Study from Bikaner district of Rajasthan, India conducted in 510 TB patients during April 2010 to January 2011 reported only 19.6% patients knew that TB is infective [23]. Possible reasons for such wide variations can be differences in population, time of conduct of study and information, education & communication activities under programme in these areas. However, our finding is similar to the findings reported from country wide data and from general population of Delhi [9] [16].

Total 88.9% participants of our study knew that TB transmits from patient to person in close contact by coughing or sneezing by patient. Available data from various states and population of India report wide variation [9] [10] [11] [13] [17] [19]. The reported range of knowledge on ‘TB transmits from patient to person in close contact by coughing or sneezing’ is 20.0% - 71.8% [11] [9]. Study on general population of Delhi reported 71.8% subjects knew coughing / sneezing as the mode of transmission [9]. Study from tribal population of Jharkhand reports 38.0% participants knew TB transmits through cough droplets [10]. 20.0% rural population of Tamil Nadu knew the mode of spread of TB [11]. A nationwide study on 198,718 participants found 55.5% knew about the correct mode of tuberculosis transmission i.e. “Through the air when coughing or sneezing” [13]. Cross sectional study conducted among 360 college students of Karnataka showed 38.9% students knew correct mode of transmission [17]. The study on TB patients in AIIMS, Rishikesh, Uttarakhand, a tertiary care centre in northern India reported only 51.5% patients knew about its causation [19].

As it is understood from previous studies, knowledge on mode of transmission of TB varies by not only by geography but also by type of participants. The knowledge on mode of transmission among rural population ranged from 20.0%-38.0% in previous studies [10-11]. Our rural participants showed even higher coverage of the knowledge on mode of transmission. The possible reasons for this difference can be local population level characteristics like education, age, & information of services of TB control programme and period of study undertaken.

About 80.3% of our participants knew that TB is treatable. This value is higher than the reported values 62.3% for rural people from Telangana [18].

In this study, 72.5% of participants knew that the diagnosis and treatment of TB are provided free of cost at government run health centres. The reported knowledge on this variable in previous studies varies from 34.0% to 91.0% from different geographical areas and population. Only 34.0% rural people from Tamil Nadu knew that treatment for TB was available free of cost [11]. Study by Muniyandi M *et al* among Saharia tribal population in Madhya Pradesh during December 2012-July 2013 reported 91.0% knew that TB diagnosis, and treatment facilities were available in both government and private hospitals [15]. Study by Shedole DT *et al* among college students of Karnataka found 71.7% participants knew about the availability of diagnostic and treatment facilities [17]. Study among young rural people of Telangana state found 89.0% knew about free diagnostic services and 85.3% free treatment services in the government setup [18]. 73.5% knew that treatment is available at government health centre free of cost [23]. Our finding is similar to findings of most of the previous studies undertaken in country.

There were two study variables which showed poor knowledge in our studied participants. About 22.3% participants of our study knew that ‘DOTS is the name of treatment for TB’. Previous studies also reported similar findings from other parts of country. A community-based cross-sectional survey carried out amongst tribal population in Madhya Pradesh reported 67.2% did not know the DOTS programme, in other words 32.8% knew that DOTS is treatment for TB [12]. Only 18.6% rural people in Telangana knew about DOTS [18]. Only 9.8% TB patients from Bikaner, Rajasthan knew about DOTS [23]. So, knowledge on DOTS was reported poor in most of the reported studies, including our study.

Second variable which reported poor knowledge was ‘*sputum is used for diagnosing TB’*. Only 58.9% participants of this study knew that the diagnosis of TB is made through examining the sputum at health centers. Kulkarni P *et al* reported 37.0% tribals knew sputum examination as method of diagnosis [10]. Rural people of Telangana showed better knowledge as 71.3% knew sputum is used for diagnosis [18]. The finding of our study is in the range of published reports.

The poor performing knowledge indicators of our study ‘*DOTS is the name of treatment of TB’* and ‘*sputum is used for diagnosing TB’* were subjected to bivariate analysis for comparing the socio-demographic factors and inequity if any (Table 2 and Table 3).

Although in our study knowledge on ‘*DOTS is the name of treatment of TB’* was poor among most of the studied socio-demographic characteristics, however proportions of participants among *SC, MBC* and *OBC* who knew ‘*DOTS is the name of treatment of TB’* were statistically significantly lower than the proportion of participants among *General* social group (p<0.05). Similarly, proportion of participants who knew ‘*DOTS is the name of treatment of TB’* among male and age groups 25 yrs and more were statistically significantly lower than the proportion among female and 15 – 24 yrs age groups respectively(p<0.05).

Proportions of participants knowing ‘*sputum is used for diagnosis of TB’* among *SC, ST*, and *OBC* were statistically significantly lower than the proportion among *General* social group (p<0.05). Similarly, proportion of participants among Hindus who knew were statistically significantly higher than the proportion among Muslims (p<0.05).

Previous published studies on socio-demographic inequity on knowledge on TB and services of TB control programme provide some data. Nationwide cross sectional survey carried out by Sreeramareddy CT *et al* found being male (aOR 1.17, 95% CIs 1.14, 1.21), being a Hindu (aOR 1.20, 95% CIs 1.14, 1.26) or Muslim (aOR 1.26, 95% CIs 1.18, 1.34), were associated with correct knowledge on tuberculosis transmission [13]. This study did not mention about socio-demographic inequity on knowledge on ‘*DOTS is the name of treatment of TB’*. Similarly study by Mukherjee S *et al* on socio-demographic factors found that the extent of knowledge about TB was more in the age group between 25 to 40 years than any other age group [14]. Study by Sharma SK *et al* found that Hindu participants had statistically significantly higher knowledge [19]. However, there is scarcity of data addressing socio-demographic inequity with respect to knowledge on ‘*DOTS is the name of treatment of TB’* and knowledge on ‘*sputum is used for diagnosis of TB’*. The present study provides this pertinent data and report socio-demographic inequity of knowledge on ‘*DOTS is the name of treatment of TB’* and knowledge on ‘*sputum is used for diagnosis of TB’*.

Multivariable logistic regression analysis found out socio-demographic inequity with knowledge on ‘*DOTS is the name of treatment of TB’* and ‘*sputum is used for diagnosis of TB’*. Being male, older age group, and belonging to *MBC, OBC, SC* & *ST* social groups are likely to be less knew on ‘*DOTS is the name of treatment of TB’*. Similarly being older age, Muslim, and belonging to *OBC, SC* & *ST* social groups were likely to be less knew on ‘*sputum is used for diagnosis of TB’*. Being male was more likely to knew ‘*sputum is used for diagnosis of TB’*.

## Conclusion

Rural population of district Jaipur, Rajasthan adequately know that the TB spreads from its patient to person in close contact through coughing and/or sneezing. They are poor in knowing that sputum is used for diagnosing the tuberculosis disease and DOTS is the name of treatment of TB. Socio-demographic inequity exists in this knowledge. People from older age groups, underprivileged social groups and minority need extra educational activities.

## Data Availability

All relevant data are within the manuscript and its Supporting Information files.

## Declarations of interest

None

## Source of funding

Study was funded by Model Rural Health Research Unit, Bhanpur Kalan, Jaipur. Unit supported the travel expenditures and provided the technical staff for data collection, and data entry.

## Author contributions

PKA, GST, MR, & MLM contributed to the study conception and design. Questionnaire was prepared by PKA, MLM, MR. Data collection was performed by PKA, MS, CRM, MRD, AJ, & BMS. Data entry and analysis was performed by PKA, MLM & BMS. The first draft of the manuscript was written by PKA and all co-authors commented on first draft of manuscript. All authors read and approved the final manuscript.

## Acknowledgement

Authors are thankful to the Dr. A.K. Sharma, Director, ICMR – National Institute for Implementation Research on Non Communicable Diseases, Jodhpur for his valuable guidance and supports.

## Notes

### Competing Interest Statement

The authors have declared no competing interest.

### Funding Statement

The author(s) received no specific funding for this work.

### Author Declarations

Institute Ethics Committee of ICMR - National Institute for Implementation Research on Non Communicable Diseases (formerly Desert Medicine Research Centre), Jodhpur

